# MyPlate, Half-Plate, and No Plate: How Visual Plate-related Dietary Benchmarks Influence Meal Composition

**DOI:** 10.1101/2021.08.05.21261632

**Authors:** Brian Wansink, Audrey Wansink

## Abstract

Can visual plate-related dietary guidance systems – such as the MyPlate guideline or the Half-Plate Rule – help people eat better when dining at home or in restaurants? To help explore this, 104 young adults completed a food diary study after having been randomly assigned to follow either 1) USDA MyPlate guidelines, 2) the Half-Plate Rule, or 3) no guidelines (control condition). Both of the visual dietary guidance systems were considered easy to understand, to follow, and left people with fewer questions about what to eat (all p<.01). Moreover, people who rated a system “easy to follow” indicated they had consumed less (meat (*r* = .268), but this was uncorrelated with fruit and vegetable intake (*r* =.092) and carbohydrate intake (*r* = .069). There are three key conclusions to these and other findings: First, the simplest guidance system may be more effective than no system. Second, even the most perfect dietary guidance system will not change behavior if the foods are not available or it is not followed. Third, guidance systems could *over-*increase the consumption of any food they specifically mention.

## Introduction

Eating balanced, nutritious meals at home is difficult for all ages – from young people who are just leaving home as well as for elderly people who are increasingly trying to stay at home as they age. Technology platforms make it possible for people to eat healthier (Win 2017; Kon, Lam, and Chan 2017), and recent trends in nutrition research have focused on how the “rules of thumb” might prove to be useful in helping individuals make better meal-related decisions whether at home or dining out (Fernández-Barrés, et al 2017). Specifically, how do different simple visual dietary guidance systems such as the MyPlate or the Half-Plate Rule influence eating behaviors compared to people who follow no eating guideline?

One useful way to address mindless eating is to provide a dietary guidance system to help people quickly determine which foods to eat in the appropriate proportion (Welsh, Davis, & Shaw, 1992). Although dietary guidance is largely available through websites and apps, people need nutrition guidelines or rules of thumb that can quickly be remembered and used in the moment (Saghafi-Asl and Vaghef-Mehrabany 2017). One example of such dietary guidance is the U.S. Dietary Guidelines. Until 2009, this guidance system was graphically represented by an image a Food Pyramid, which was referred to as “MyPyramid.” In 2009, this pyramid was modified into the form of a plate that was proportionally divided into four quarters that represented components of grains, proteins, fruits, and vegetables (along with a serving dairy on the side of the plate, which was represented as a glass of milk). This system became quickly adopted, especially by those who had been well-educated, had children, or who believed that this guideline would work for their health (Wansink & Kranz, 2013).

Since its introduction, this MyPlate icon was widely used by the government to represent the more lengthy and complete 149-page dietary guidelines. Considering that nutrition information sometimes seem too complex to be an actionable (Cowburn & Stockley, 2005; Grunert & Wills, 2007), MyPlate was designed to offer a quick visual and actionable summary and easy to follow benchmark. In doing so, it was intended to prompt diners to think about eating more balanced meals, such as ones which included more fruits, vegetables, and whole grains (Bachman, Reedy, Subar, & Krebs-Smith, 2008; Post, Haven, & Maniscalco, 2011). Yet there is a concern that some that people may not be following any dietary guidance system (Kon, Lam, and Chan 2017). For example, if they lack motivation and nutrition knowledge, they may not understand how to categorize their food into the recommended components--grain, protein, fruits and vegetables and dairy (Wansink & Kranz, 2013). Moreover, even when users of the MyPlate or MyPyramid do understand a rule, issues like individual food preferences and tastes could influence whether they actually followed these guidelines. Therefore, it is necessary to examine whether people *understand* the guidelines but also whether they *follow* them.

A second rule-of-thumb dietary guidance system – the Half-Plate Rule – is complementary to the MyPlate approach, but it is a more basic system that has been utilized in school cafeterias, dining halls, and grocery stores (Wegman’s grocery stores refers to it as Half-Plate “Healthy”). The Half-Plate Rule that suggests whenever a person makes an eating decision (such as what to order or what to serve themselves), they should aim for half of their plate to be filled vegetables, fruits, or salad, while the other half should contain a reasonable balance of anything else (Wansink 2014). The key question is: Will such guidance systems provide people with the confidence to eat better and lead them to consume less food and have more relatively balanced nutrition meals? (Guthrie, Mancino, & Lin, 2015).

When interventions and instructional advice are given, it is often believed that the simpler the approach, the higher the adherence. For instance, simplicity and unambiguousness are suggested to be two reasons why diets achieve quick success with some people (Sunstein, 2016). Over-explicitly raise awareness of the variety of foods, might people some to eat more of those foods than they otherwise would. Categorization research has shown that the more categories are presented to persons, the more foods people take (Simonson, 1999).

This research aims at determining how dietary guidance systems might influence diners to consciously eat better. By doing so, this offers an important way that Smart Homes, apps, and technological platform could also communicate or track eating behavior in a helpful way. In doing so, the results of this study can also give health professionals and public health officials insights into the types of guidance that they can use to more effectively influence eating behaviors. Such findings would also contribute to the meaningful debate on whether it is more effective to about how to best present dietary guidance information

## Materials & Method

To initially determine the effectiveness of dietary guidance systems, university students and staff were offered extra course credit if they agreed to be involved in an eating study while during a four-day holiday break. In this IRB-approved study, these individuals were randomly divided into one of three conditions. One group of participants was asked to follow the MyPlate guide system recommended by the USDA. Another group was asked to follow the Half-Plate Rule. The third group was asked to eat as they normally would (control condition). These instructions were briefly summarized on a single page of paper, which also included a graphic icon of the dietary guidance system (a MyPlate divided into four or a MyPlate divided into two).

Those people in the MyPlate guidelines condition were presented the MyPlate icon, and they were advised to have balanced amounts of fruits, vegetables, grains, proteins, and dairy for lunches and dinners. MyPlate guidelines has been adopted by US government since 2009. Those people in the Half-Plate Rule condition were shown a graphic of the Half-Plate, and they were advised to fill half of their dinner plates filled with fruits, vegetables and salad and the other half can include foods they wish. They were also told consumption amount was not an issue and they could freely refill their plates, but then still needed to follow this Half-Plate Rule. Those people in the third group were in the control condition, and they were given no guidance or rule as to what to eat.

After finishing their meal under these randomly assigned conditions, participants answered a one-page survey questionnaire that had been sent home with them. Questions were asked using a 9-point scale (1 = Strongly Disagree; 9 = Strongly Agree). The first three questions asked participants to rate how easy it was for them to understand the assigned rule and to follow it, and whether they had any questions on it. After that, participants were asked to rate questions regarding their eating behaviors—such as “I ate healthier than usual”, “I ate less food than usual”, “I ate more fruits and vegetables”, “I ate less dairy than usual” and so on. The final part of the survey required participants to estimate the total calories that they ate, and to answer how many different foods they ingested, as well the snacking calories. They were told to use this rule at any or all meals they consumed during that four-day period, but they were to answer the survey immediately after one specific evening dinner meal that was not a special holiday meal or celebration meal. They could select whichever dinner they wished.

Among those participants who initially agreed to be involved in the study, 69 analyzed an at-home meal and provided their answers in time for analysis. The remainder of individuals analyzed meals at restaurants. Since these responses were not relevant for our investigation of at-home eating behaviors, they were separated and excluded from any of these analyses.

The foods that respondents had reported eating were categorized for analysis, but because many people did not include specific enough indications of their serving size, these data were not ultimately analyzed. Similarly, although self-reported measures of calorie intake were asked in the questionnaire, such measures can be highly variant and inaccurate. Although one way to analyze this data is to exclude outliers, it was believed to be more prudent to include summary measures of them in the tables, but not to emphasize them in the discussion of the analyses.

The analyses were conducted using the Wizard (Version 1.9.48) Statistical Analysis software to assess the mean value of rating scores and self-reported calorie intakes. To test the difference between the individual conditions, we performed independent t-tests. The Pearson correlation test was also conducted to examine if there is any relationship among variable variances.

## Results

As shown in Table 1, when comparing participants who used a guidance system such as the Half-Plate or MyPlate guidelines versus those following no dietary guidance system (control condition), it is found that using dietary guideline systems generally led participants to have fewer questions about what to eat (2.36 vs. 3.76 vs. 4.13; F _(2, 65)_ =3.80, *p*=.027). Of the two dietary guidance systems, those using the Half-Plate Rule reported that it was easier to follow (6.59 vs. 5.12; t _(45)_ =2.37, *p*=.022) and easier to understand (8.62 vs. 7-68; t _(45)_ =2.02, *p*=.049) than the MyPlate guidelines.

**Table 1.** How Dietary Guidance Systems Influence At-Home Eating Behaviors.

(Standard Deviations and P-values are in Parenthesis)
|  | Half-Plate<br>Rule<br>n=35 | MyPlate<br>n=35 | Control<br>n=34 | F-test<br>(2,103)<br>(p-value) |
| --- | --- | --- | --- | --- |
| **Understandability** |  |  |  |  |
| It was easy to follow this rule<sup>1</sup> | 5.41<br>(0.74) | 5.03<br>(0.69) | 6.81<br>(0.65) | 7.61***<br>(0.001) |
| It was easy to understand this rule<sup>1</sup> | 6.85<br>(0.75) | 7.67<br>(0.50) | 8.31<br>(0.26) | 3.047***<br>(0.001) |
| I had questions about it<sup>1</sup> | 4.50<br>(0.89) | 4.03<br>(0.80) | 2.64<br>(0.65) | 6.31***<br>(0.003) |
| **Targeted Eating Behaviors** |  |  |  |  |
| I ate healthier than usual<sup>1</sup> | 5.53<br>(0.68) | 5.58<br>(0.56) | 5.83<br>(0.77) | 0.024<br>(0.89) |
| I ate less unhealthy than usual<sup>1</sup> | 5.44<br>(0.67) | 5.0<br>(0.68) | 4.61<br>(0.76) | 1.48<br>(.249) |
| I ate more fruits, vegetables, and<br>salad<sup>1</sup> | 6.12<br>(0.61) | 6.00<br>(0.73) | 6.25<br>(0.77) | 3.085<br>(0.878) |
| I ate less carbs than usual<sup>1</sup> | 4.79<br>(0.64) | 4.75<br>(0.71) | 5.5<br>(0.82) | 1.383<br>(0.256) |
| I ate less meat than usual<sup>1</sup> | 4.71<br>(0.69) | 4.25<br>(0.68) | 5.20<br>(0.79) | 1.785<br>(0.173) |
| I ate less dairy than usual<sup>1</sup> | 4.65<br>(0.77) | 3.6<br>(0.76) | 4.9<br>(0.84) | 3.722**<br>(0.030) |
| I ate fewer desserts than usual<sup>1</sup> | 5.61<br>(0.81) | 5.53<br>(0.97) | 6.33<br>(0.85) | 1.05<br>(0.357) |
| I snacked less than usual<sup>1</sup> | 5.59<br>(0.71) | 5.56<br>(0.65) | 4.69<br>(0.69) | 2.284<br>(0.107) |

<sup>1</sup>Note: \*p< .10, \*\*p<.05, \*\*\* p<.01; Rating scale: 1 = Strongly Disagree to 9 = Strongly Agree; Cell sizes vary by question since not all questions were answered by all respondents

**Table 2.**
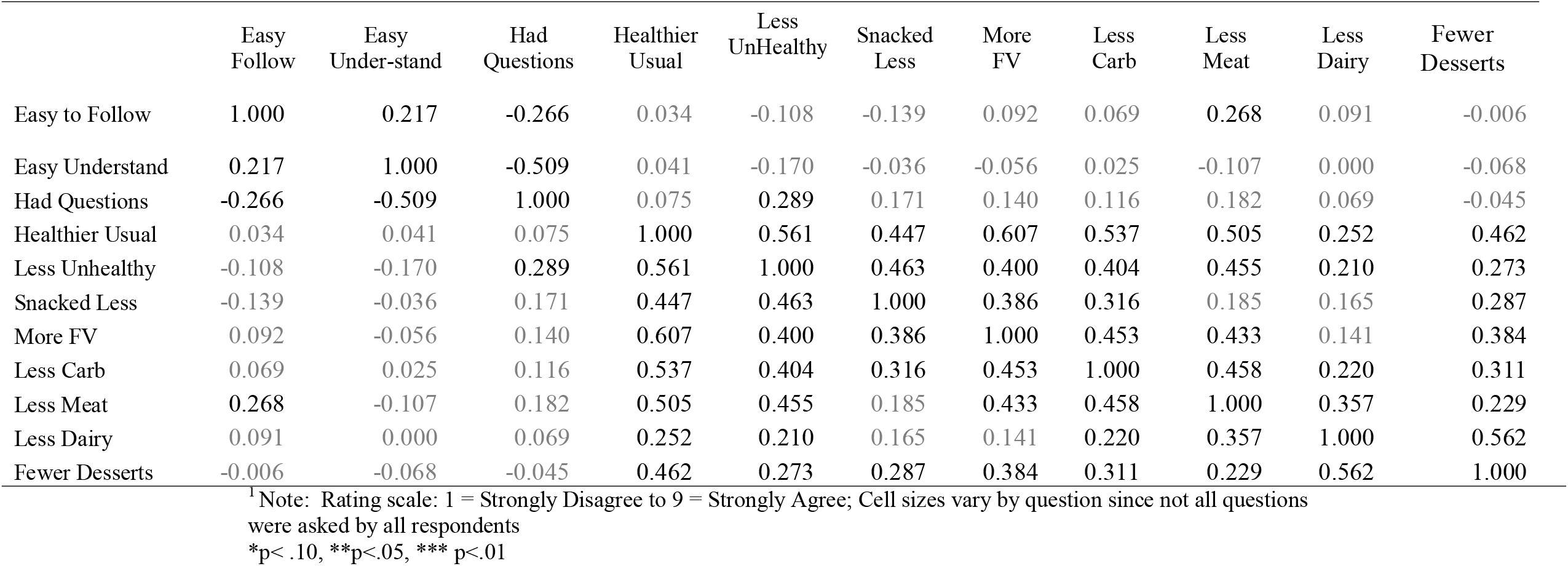
Pearson’s Correlations between Eating Behaviors.

|  | Easy Follow | Easy Under-stand | Had Questions | Healthier Usual | Less UnHealthy | Snacked Less | More FV | Less Carb | Less Meat | Less Dairy | Fewer Desserts |
| --- | --- | --- | --- | --- | --- | --- | --- | --- | --- | --- | --- |
| Easy to Follow | 1.000 | 0.217 | -0.266 | 0.034 | -0.108 | -0.139 | 0.092 | 0.069 | 0.268 | 0.091 | -0.006 |
| Easy Understand | 0.217 | 1.000 | -0.509 | 0.041 | -0.170 | -0.036 | -0.056 | 0.025 | -0.107 | 0.000 | -0.068 |
| Had Questions | -0.266 | -0.509 | 1.000 | 0.075 | 0.289 | 0.171 | 0.140 | 0.116 | 0.182 | 0.069 | -0.045 |
| Healthier Usual | 0.034 | 0.041 | 0.075 | 1.000 | 0.561 | 0.447 | 0.607 | 0.537 | 0.505 | 0.252 | 0.462 |
| Less Unhealthy | -0.108 | -0.170 | 0.289 | 0.561 | 1.000 | 0.463 | 0.400 | 0.404 | 0.455 | 0.210 | 0.273 |
| Snacked Less | -0.139 | -0.036 | 0.171 | 0.447 | 0.463 | 1.000 | 0.386 | 0.316 | 0.185 | 0.165 | 0.287 |
| More FV | 0.092 | -0.056 | 0.140 | 0.607 | 0.400 | 0.386 | 1.000 | 0.453 | 0.433 | 0.141 | 0.384 |
| Less Carb | 0.069 | 0.025 | 0.116 | 0.537 | 0.404 | 0.316 | 0.453 | 1.000 | 0.458 | 0.220 | 0.311 |
| Less Meat | 0.268 | -0.107 | 0.182 | 0.505 | 0.455 | 0.185 | 0.433 | 0.458 | 1.000 | 0.357 | 0.229 |
| Less Dairy | 0.091 | 0.000 | 0.069 | 0.252 | 0.210 | 0.165 | 0.141 | 0.220 | 0.357 | 1.000 | 0.562 |
| Fewer Desserts | -0.006 | -0.068 | -0.045 | 0.462 | 0.273 | 0.287 | 0.384 | 0.311 | 0.229 | 0.562 | 1.000 |

<sup>1</sup> Note: Rating scale: 1 = Strongly Disagree to 9 = Strongly Agree; Cell sizes vary by question since not all questions were asked by all respondents
\*p< .10, \*\*p<.05, \*\*\* p<.01

People who believed that their dietary guidance system was easy to follow tended to report eating slightly better. There was a significant correlation between a system being easy to follow and a person eating less meat (*r*=.268; *p*<.05). In contrast, having an easy to follow system had no impact on whether a person reported eating healthier eating (*r*=.034; *p*>.05), eating more fruits and vegetables (*r*=.092; *p*>.05), or eating less carbohydrates (*r*=.069; *p*>.05).

Yet even though there were few positive correlations between being easy to follow and eating better, neither of the two individual guidance systems translated into people reporting they ate better. People who followed either of the two dietary guidance systems did not report that that ate any healthier (*p*>.20) than those following no system. As Table 1 indicates, the one exception was that those people who were given MyPlate guidelines ate the most dairy (compared to usual), and those following the MyPlate guidelines ate the most dairy (3.60 vs. 4.65 and 4.90; F _(2,103)_ = 3.722, *p*=.03).

Following the study, debriefing interviews were conducted. They revealed an unexpected explanation as to why dietary guidance systems improved understanding of how to eat better, but then had little impact on actual eating behavior. These interviews indicated that many of these meals did not have a wide variety of fruits and vegetables available. As a result, there was not the opportunity to substitute them for the carbohydrates, grains, or meat and protein in the way suggested by the guidelines. This made following either the MyPlate Guidelines or the Half-Plate Rule very difficult. That is, although they could control what they put on their plate, they were limited by what was put on the table in front of them.

## Discussion

One powerful advantage of dietary guidance systems is that they may be one way give an elderly person more confidence in better knowing about how they can eat healthier (Kolodinsky, Harvey-Berino, Berlin, Johnson, & Reynolds, 2007). Dietary guidance systems can help people move from mindless eating to more mindful eating (Fernández-Barrés, et al 2017). The more simple the guidance system, the more confident a person feels when making healthy food choices (Just, et al, 2007). That may be one strong reason as to why some very simple—albeit controversial—systems, such as low carbohydrate diets have been proven to be very popular for at least a brief period of time (Astrup, Larsen, & Harper, 2004; Dansinger, Gleason, Griffith, Selker, & Schaefer, 2005; Malik & Hu, 2007). A simple rule as to what to eat— such as the MyPlate or the Half-Plate Rule – can provide a bounded direction that encourages people to eat a wider variety of healthier foods.

Between these two dietary guidance systems, the Half-Plate Rule, which unambiguously divides food into only two categories, was rated as easier to follow and understand than the MyPlate guidelines, which divides food into five different categories. Moreover, it was generally found that the more understandable a dietary guidance system was to a person, the less meat, less dairy, and less dessert they consumed.

Although both dietary guidance systems left people with fewer questions on what they should eat, neither was particularly effective at dramatically changing how people ate. That is, they believed the dietary guidance systems were easier to understand, yet they did not always claim to eat better. A follow-up series of discussions with these individuals indicated that there were not always enough fruits and vegetables available on the table for them to eat in the way suggested by the guidelines. Using the Half-Plate Rule can even be difficult if there is only one fruit or vegetable available. Perhaps Smart Homes could facilitate the availability of fruits and vegetables through technology or applications that helped monitor food inventory, or which provided recipe ideas would help reduce this gap between understanding what to eat and doing it.

It was surprising to see, however, that the MyPlate guidelines led people to eat more dairy than they otherwise would. It might be that certain individuals who typically do not consume dairy within their normal meal were guided or reminded by the MyPlate guidelines to consider having dairy in their meals. This can be potentially advantageous in situations where people need to consume more calcium such elderly people who are the focus of some of many promising Smart Home projects. It can also be advantages for children, people with very active or low iron level, and people who require high nutrition food portion for special hospital treatment. However, it may not be as advantageous to other groups of people who are already consuming too many calories, and who may consume these products in the form of fattier dairy products (such as butter, whole milk, ice cream, and so on).

### Limitations and Future Research

One of the strengths of this study was that the experiment was conducted in natural at-home dinners where participants could freely use a dietary guideline to decide what they ate. Yet, as an initial study in this area, there are several limitations to this study. First, because of the wide variation in eating conditions people experience, it would have been useful to conduct this study with a larger sample size that consisted of people at different ages and with different cooking capacities.

A second limitation of this study is that most people do not regularly keep a diet food diary. As a result, knowing that you will have to write what you will be eating may alter the way a person eats a meal. Future studies that can more carefully observe what people serve and how much they consume during meals, and it could also investigate how these variations might change across different meals. For instance, it might be expected that a guidance system might have less influence over a breakfast or lunch than it does a dinner (where more food is available). across meal occasions. Furthermore, the variance in demographic of our participants was not large in terms of education or economic background. Past research on MyPlate, for example, had shown the people who most quickly adapted to my plate with those who were the most educated and those who were the most attuned to their own dietary pattern (Wansink & Kranz, 2013).

### Implications

There are three key conclusions and related implications to this study:

- First, a simpler guidance system may be more effective than a complex one. When analyzing the people who rated the guidance system as easy to use versus easy to understand, it was found the simpler the system is, the easier it was understood and the more correlated it was with selected healthier eating behaviors.
- Second, even the most perfect dietary guidance system will not change behavior if the foods are not available. No dietary system will change behavior if only hamburger and chips are available for lunch. What such a system can do, is to eventually encourage greater variety to be added that will balance out the meal.
- Third, guidance systems may increase—for better or for worse—the consumption of any food they specifically mention and highlight and even they may decrease the consumption of that food. Raising the awareness of food that a person may not have otherwise eaten or eat in significant quantitative such as dairy, meat or starches could unconsciously influence that person when he adheres to the systems specifically mentioned.

Dietary guidance systems such as MyPlate have been shown to be more understandable compared to other systems such as MyPyramid (Krebs-Smith, Guenther, Subar, Kirkpatrick, & Dodd, 2010; Wansink 2008). In this study, the Half-Plate Rule and MyPlate guidelines are much more efficient in giving people the confidence in what they choose to eat and in part, eating a better-balanced meal than when using no system at all. In general, these dietary systems may be effective in helping people eat more mindfully.

## Conclusions

Dietary guidance systems are useful ways to encourage more mindful eating. Moreover, they can be easily modified to be used with apps and monitoring devices, or even in basic ways that are as simple as a reminder icon or graphic. Given the pressures on many people’s cognitive load, the simplicity or these apps or monitoring devices is critical. The simpler they are, the more flexible they are, and the more effective they will be. For instance, telling a person that half their plate should be fruit, veggies, and salad would be one way to provide to them the flexibility to lately eat what they want but to balance it in the simple ways in which otherwise may not have been done.

## Data Availability

Data will be available upon publication

